# Applying SARS-CoV-2 serological testing to understand the effectiveness of local infection control measures for COVID-19 infection in Victoria, Australia 2020

**DOI:** 10.1101/2025.08.22.25334274

**Authors:** K.A. Bond, V.K.Y. Leung, S. Lim, D.F. Johnson, B Lin, A. Harding, C. Marshall, B.P. Howden, D.A. Williamson, E Tippett

## Abstract

The availability of early serological assays for COVID-19 infection allows investigation of aspects of SARS-CoV-2 transmission which may not be evident on PCR testing. Here we describe two serological surveys in different settings in 2020 which assess the effectiveness of local infection prevention practices: i) a retrospective cohort of household contacts previously quarantined; and ii) health care workers in a tertiary hospital. Serological testing of household contacts did not diagnose any additional cases to that which had been notified by testing symptomatic contacts with PCR. The secondary household attack rate for these 53 contacts from 42 households was 36.2% (95% CI: 37.4, 74.5%). Sero-positivity in health care workers increased from 0.5% in May 2020 to 3.8-5.5% in August to October 2020. Multivariate analysis demonstrated that male HCW were less likely to be sero-positive (OR 0.19 95% CI 0.03-0.7, p=0.04), while there was an increased risk of sero-positivity associated with nursing staff (OR 8.1 95% CI 1.7-145.6, p=0.04), staff who experience anosmia (OR 22.7, 95% CI 5.4-112.4, p < 0.001); and those caring for COVID-19 patients (OR 4.1 95% CI 1.8-10.5, p=0.01). In this low prevalence setting with rapid access to PCR testing, serological testing provided a small additional benefit to PCR testing alone. Measures to detect COVID-19 cases in household contacts were found to be sufficient, while infection control measures during a period with a high burden of cases in a tertiary hospital were found to be inadequate to prevent COVID-19 infections in health care workers.

## Introduction

Understanding the virology and transmission dynamics of SARS-CoV-2 is key to appropriately tailor public health community and health care responses. Fortunately the early development of robust molecular assays has supported the test-trace-isolate strategy that has been critical in implementing Australia’s strategy to maintain zero community transmission until the widespread availability of COVID-19 vaccines(1). However PCR testing alone is unlikely to provide all the information we require to fully understand COVID-19 transmission dynamics, vulnerability to infection and key population risk factors, and is more labour intensive and expensive to test at scale. By mid 2020, early laboratory-based commercial serological assays have been developed and become commercially available for use, allowing application locally to explore some of these knowledge deficits.

Following the implementation of a series of public health and social restrictions, by May 2020 the majority of Australian states experienced a very few incident cases of COVID-19 infection(2). Victoria, Australia’s second most populous state, experienced the greatest burden of COVID-19 in 2020. This included a ‘Wave 1’ of local COVID-19 cases in March and April, which was driven by overseas travellers and their direct contacts. Following a series of staged restrictions on public activities, incident cases approached zero by May. A ‘wave 2’ of COVID-19 infection was experienced from mid-June to early October, after the relaxation of some public health measures and a large community outbreak (~18,000 cases, peak incidence ~550/100,000 per year, ~0.3% Victorian population) which was traced to an introduction from returning citizens undertaking quarantine at central hotels(2,3). This setting provides a useful environment in which to study aspects of community and health care related COVID-19 transmission, where contacts are likely to have a single exposure to COVID-19 and the subsequent effects of public health infection control measures can be observed.

To ensure minimal transmission of COVID-19, finding all cases of COVID-19 is critical to allow complete contact tracing and isolation of cases and contacts. Serological testing can be helpful in diagnosing PCR negative or asymptomatic cases which may be integral to transmission chains. Additionally, serological testing can also improve our understanding of the limitations of PCR, to ensure PCR is utilised in the most resource effective manner. In this study, our objective was to rapidly implement SARS-Co-2 serological testing in 2020 to understand COVID-19 transmission in two local settings, and provide early feedback on the effectiveness of the infection control strategies employed locally.

The first setting was that of households required to quarantine following the diagnosis of COVID-19 in one or more household members. We aimed to determine whether the current policy of only testing symptomatic household contacts with PCR missed any transmission events for contacts in home quarantine. As a secondary objective we aimed to determine the COVID-19 secondary attack rates for these households, and whether any epidemiological risk factors were associated with an increased likelihood of transmission.

The second setting was that of a tertiary hospital, where staff in personalised protective equipment (PPE) were caring for COVID-19 infected patients on cohorted wards. We aimed to determine the sero-prevalence of COVID-19 in this staff cohort following wave one and wave two of community COVID-19 infections in Victoria. As a secondary objective we aimed to identify whether any clinical or epidemiological risk factor was associated with an increased likelihood of staff sero-positivity.

## Methods

This study involves two study populations: i) quarantined household contacts of COVID-19 cases; and ii) health care workers, recruited from a single tertiary hospital, Royal Melbourne Hospital (RMH), Victoria. Recruitment occurred from May to December 2020, following wave 1 and during wave 2 of localised COVID-19 outbreaks in Victoria, Australia (Figure 1). RMH is a university-affiliated tertiary hospital with 550 beds on a main campus (Campus 1), and a further 150 lower acuity geriatric and rehabilitation beds on 6 wards at a secondary campus (Campus 2), with approximately 10,000 employees in total. All pathology testing is undertaken in a National Association of Testing Authorities (NATA) accredited laboratory located on Campus 1.

**Figure 1:**
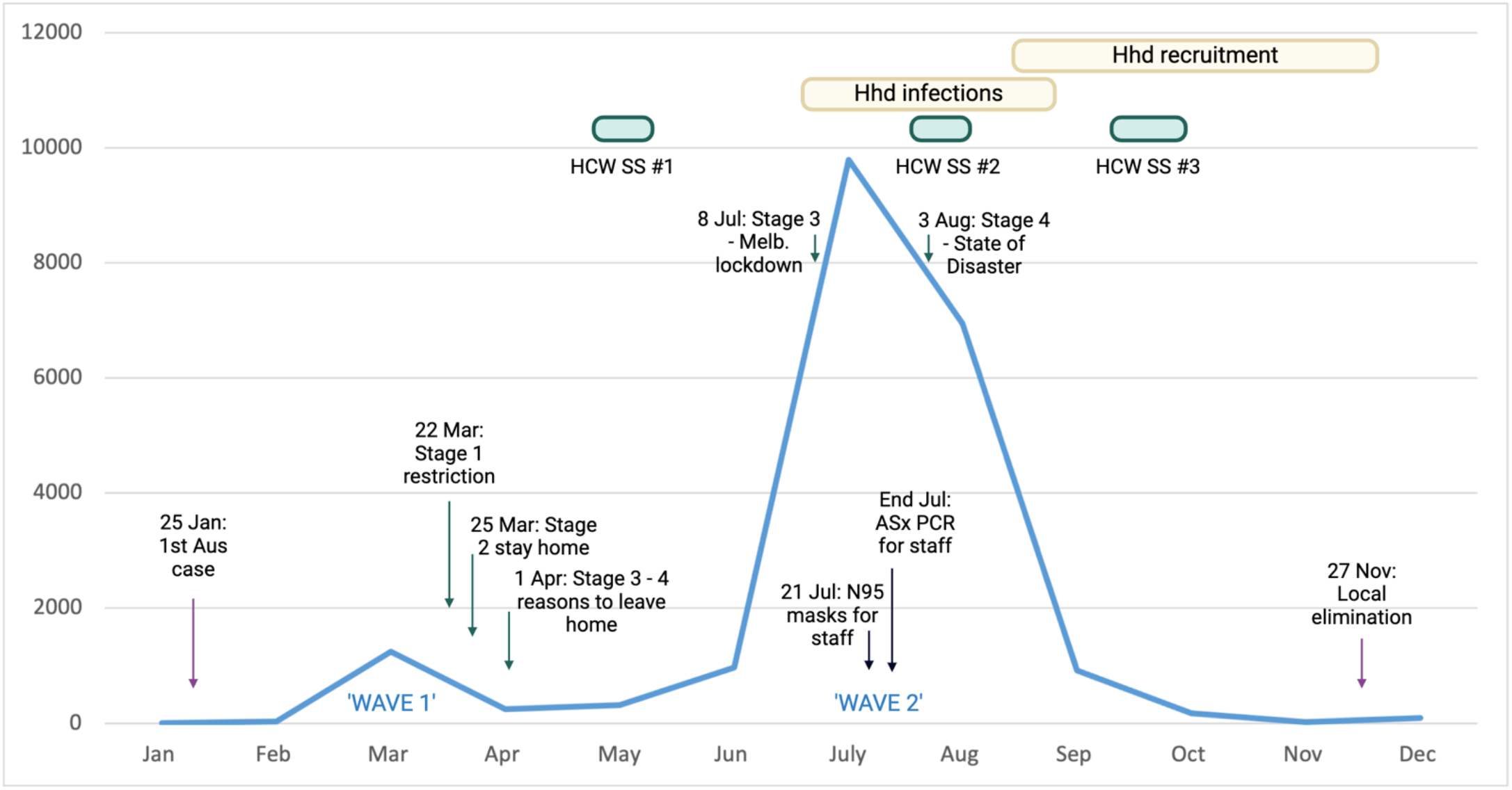
Count of COVID-19 notified cases 2020, Victoria. Monthly case count data from National Notifiable Surveillance System (https://nindss.health.gov.au/pbi-dashboard/) Stages of public health restriction of activity. Stage 1 - shutdown all non-essential activity. Stage 2 - Stay home from work and school; Stage 3 - only 4 reasons to leave home (essential workers, care or care giving, daily exercise, or buying food or other essentials); Stage 4 - state of disaster, police have greater power to enforce public health orders. SS = Sero-survey; Hhd = Household; Melb = Melbourne; ASx = asymptomatic; Jan = January; Mar = March; Apr = April, July = July; Aug = August; Nov = November

### Study setting I)

During the study period, public health regulations required all persons infected with COVID-19 to isolate in their home for 14 days following symptom onset. Any close contacts, including any household contacts, were also required to quarantine for 14 days. Co-habiting index cases and close contacts were advised to separate as much as possible within the household. Public health officers conducted telephone-based health checks for cases and contacts, with SARS-CoV-2 PCR testing undertaken on a combined deep nose and throat swab for any contacts who became symptomatic. Any contacts who subsequently developed COVID-19 infection were then isolated for a further 14 days from their symptom onset.

### Population I) Household contacts in home quarantine

Household contacts were retrospectively recruited from 25^th^ August to 1^st^ December 2020 as part of a larger prospective observational study which recruited confirmed cases of COVID-19 infection for serological testing at baseline (recruitment), 3, 6 and 12 months following infection. Adult COVID-19 cases confirmed through the study laboratory within the previous 12 weeks (9^th^ June to 8^th^ September), and who lived in a geographical region serviced by the RMH@Home nursing service, were offered enrolment in the primary study via telephone. Terminally ill patients or those with significant cognitive impairment were excluded. Enrolled COVID-19 cases (index case) were asked if there were any household contacts (those staying overnight in the household within 48 hours prior to symptom onset or within the 14 day isolation period) who would like to take part in the Household contact sub-study. Contacts under 18 years were not eligible for enrolment. Contacts were eligible for enrolment in the household sub-study whether they had or had not been symptomatic during quarantine, had or had not been diagnosed with COVID-19 at time of enrolment, and whether or not all members of the household enrolled. COVID-19 diagnosis in household contacts was confirmed by review of accredited laboratory PCR report or evidence of public health COVID-19 notification documentation.

Research investigators collected retrospective clinical and household information from COVID-19 index cases and household contacts via both telephone and in-person interviews at time of enrolment. A dedicated study nurse visited household contacts to collect serum for SARS-CoV-2 serology from all study participants at three months following infection onset in the household index COVID-19 case. On review of data for the household, the primary case was assigned to the case with first date of onset of symptoms (whether initially the index case or contact). Any other cases in the household were considered co-primary cases if symptom onset was within 1 day of the primary case, and contacts if symptoms began 2 or more days after the primary case.

### Study setting II)

At RMH, all COVID-19 infected patients were managed in a single room if possible or otherwise were cohorted and managed separately to non-infected patients by dedicated staff wearing PPE. Any COVID-19 infected staff were isolated at home for 10 days or more following symptom onset, and any staff exposed to a COVID-19 infected patient or staff without PPE (≥ 15 minutes of face-to-face contact or ≥ 2 hours in a shared space in the 48 hours prior to case symptom onset) were considered close contacts and quarantined for 14 days, according to state public health guidelines. All COVID-19 infected staff were interviewed by an infection prevention clinical nurse consultant to discuss risk factors for acquisition and to enable contact tracing. Outbreaks, (two or more epidemiologically linked staff and/or patients), were managed by a local hospital incident management team. PPE consisted of gloves, gowns, face shields and face masks. In line with national guidelines surgical masks were worn in the beginning of the pandemic, however RMH instituted a change to N95 (or P2) on 21 July 2020, in response to rising staff and patient COVID-19 infections.

The number of COVID-19 infected RMH inpatients increased significantly from early July 2020, mirroring a rise in community cases associated with wave 2 (Figure 1). A large COVID-19 outbreak involving both staff and patients was managed at Campus 2 in early July (4). Peak daily numbers of patients managed with COVID-19 at campus 2 was 60, with 107 of 262 (40.8%) staff COVID-19 infections between 1 July and 31 August registered to staff predominantly working at Campus 2, despite staff at this site constituting ~ 10% of the overall staff workforce. In contrast Campus 1 managed multiple smaller outbreaks across a larger number of wards. From late July, staff working on wards managing suspected or confirmed COVID-19 patients were offered weekly asymptomatic combined nose/throat PCR testing to detect infections early. Dedicated staff sample collection clinics were located at both Campus 1 and 2, with a median turn-around time for health care worker test results of 20.2 hours (IQR 11.4-29.1hrs).

### Population II) Health care workers in a hospital setting

RMH staff were offered optional COVID-19 serological testing at 3 time points in 2020, supported by dedicated staff sample collection clinics. Sero-survey 1 was offered to all staff over a 2-week period in May 2020 on Campus 1, a combined deep nose and throat (N/T) swab for PCR testing was collected in addition to serology for all staff. Sero-survey 2 was undertaken over a 4-week period in August at Campus 2, after weekly asymptomatic PCR screening on N/T swab became recommended (but not mandatory) for all staff working with COVID-19 patients at the end of July. Sero-survey 3 was undertaken over a 4-week period in October at Campus 1, weekly PCR surveillance remained in place for staff working on COVID-19 wards on either a N/T swab or saliva sample. Sero-surveys were advertised to staff via posters in staff COVID-19 clinics, emails to managers, and posts on RMH social media platforms.

General clinical and epidemiological information as well as information on COVID-19 exposure events was captured for sero-surveys 2 and 3. Staff participants entered their own data directly into a REDCap web-based questionnaire on personal electronic devices after provision of a study specific URL or QR code. COVID-19 severity status was based on the World Health Organisation (WHO) severity scale(5). Medical co-morbidities were those defined as those outlined in the WHO clinical management of COVID-19. Staff were provided their serology results via an RMH dedicated patient smart device application, which is in routine clinical use for all RMH patients and for which pathology test results are routinely displayed.

### SARS-CoV-2 testing

All study samples were tested in the RMH diagnostic laboratory with accredited commercial assays. SARS-CoV-2 RNA was detected by at least two of three assays, the Coronavirus Typing assay (AusDiagnostics, Mascot, Australia), Respiratory Pathogens 12-well assay (AusDiagnostics, Mascot, Australia), the Xpert® Xpress SARS-CoV-2 (Cepheid, Sunnyvale, USA) or an in-house real-time assay at the reference laboratory, using previously published primers(6–8). All initial positive results were confirmed by re-extraction from the primary sample and amplification in at least one of the remaining three assays.

Serum samples were screened with two chemi-luminescent laboratory assays, the Abbott Architect SARS-CoV-2 IgG and the DiaSorin Liaison SARS-CoV-2 S1/S2 IgG, according to the manufacturer’s instructions for use (9,10). The Abbott assays detects IgG antibodies targeted against the nucleocapsid (N) protein, while the DiaSorin assay detects IgG antibodies against the spike protein (S1/S2 subunits).

Samples testing negative in both the Abbott and DiaSorin assays were considered negative. Any sample collected prior to June 2020 (HCW initial sero-survery, May 2020) testing positive in either or both of the Abbott or DiaSorin assays was confirmed via an in-house micro-neutralisation assay at the state reference laboratory, as previously described (18–21). From June 2020 onwards, following further in-house verification of the Abbott and DiaSorin assays(11), any sample testing positive in both assays was considered positive. Given the low positive predictive value of both assays due to the extremely low prevalence of COVID-19 in the community (< 0.6% in all study periods)(2), any sample testing positive in only one assay was sent to the state reference laboratory for confirmation. Confirmation included algorithmic testing involving the Roche Cobas Elecsys Anti-SARS-CoV-2 Wantai total antibody assay and GenScript SARS-CoV-2 Surrogate Virus Neutralisation Test (sVNT), with reference laboratory results considered as final results (whether positive or negative)(12)).

### Data management and analysis

Clinical and epidemiological participant data were captured on REDCap (Research Electronic Data Capture), in dedicated case report forms for each study population(13). COVID-19 results were extracted from the laboratory information system, and staff COVID-19 status and results of external PCR testing was extracted from the Infection Prevention and Surveillance Service COVID-19 staff database (also REDCap based). Fisher’s exact test (categorical variables) and Wilcoxon rank sum test (continuous variables) were used to compare demographic characteristics by COVID status. Univariate and multivariate regression models were run, with contact status (secondary case or contact) or COVID status (positive or negative) as the outcome, and two tailed p values reported. Data were analysed in MicroSoft Excel and R version (Version 4.1.2 and 4.2).

### Ethics approval

This study was approved by the Melbourne Health Human Research Ethics Committee as part of two protocols, 2020.207/66341 and 2020/179/68355.

## Results

### Population I) Household contacts in home quarantine

Of 335 positive COVID-19 diagnoses made through the study laboratory in the study period (infections diagnosed between 9^th^ June to 8^th^ September), 209 index cases were considered eligible for enrolment in the prospective COVID-19 serological study. Of the 85 participants recruited to the parent study, 42 recruited at least one contact into the household contact sub-study of which 23 (55%) recruited all household members (Supp Fig 1). Of the 19 households with partial recruitment, 6 (32%) included households where the only missing members were children, the remaining 13 households were missing a single adult household contact. Including all households, median size was 3 (IQR 2-4). There were 53 household contacts in total, of which 23 were known to be COVID-19 positive at enrolment, while there were 30 contacts from 20 households who had either not undergone COVID-19 PCR testing or tested negative at time of recruitment (Supplementary Table 1). These contacts with unknown status were more likely to live in larger households (households with > 2 people), compared to contacts known to be COVID-19 positive at recruitment (22/30 (73%) versus 9/23 (39%), respectively, p value < 0.01). The majority of index cases recruited to the study were HCW (31/42, 73.8%), whereas only 10 of 53 contacts (18.9%) were HCW (p < 0.001). The majority of COVID-19 infections in all groups were mild, with 5 index cases and 1 contact hospitalised.

Serological testing failed to detect any cases of COVID-19 that were not previously notified, with none of the 30 household contacts with an unknown COVID-19 status testing sero-positive at 3 months. The majority of index case and known positive COVID-19 contacts tested were sero-positive at 3 months (39/42, 93% and 39/42, 87%; respectively).

On review of epidemiological data, 6 contacts were considered to be co-primary cases, leaving 47 total contacts for analysis. The overall secondary attack rate (SAR) for household contracts was 36.2% (95% CI: 37.4, 74.5%), which did not change significantly if only households with full recruitment were enrolled (SAR 35.7%). Estimated SAR and odds for transmission based on clinical and epidemiological features are present in Table 1. The only variable associated with a significantly increased odds of COVID-19 transmission was sharing a bedroom with a primary case (OR 4.44, 95% CI 1.18-18.4, p 0.03), which remained significant on multivariate analysis (OR 7.46 95% CI 1.34-53.3, p 0.03) (Supplementary Table 2).

**Table 1:**
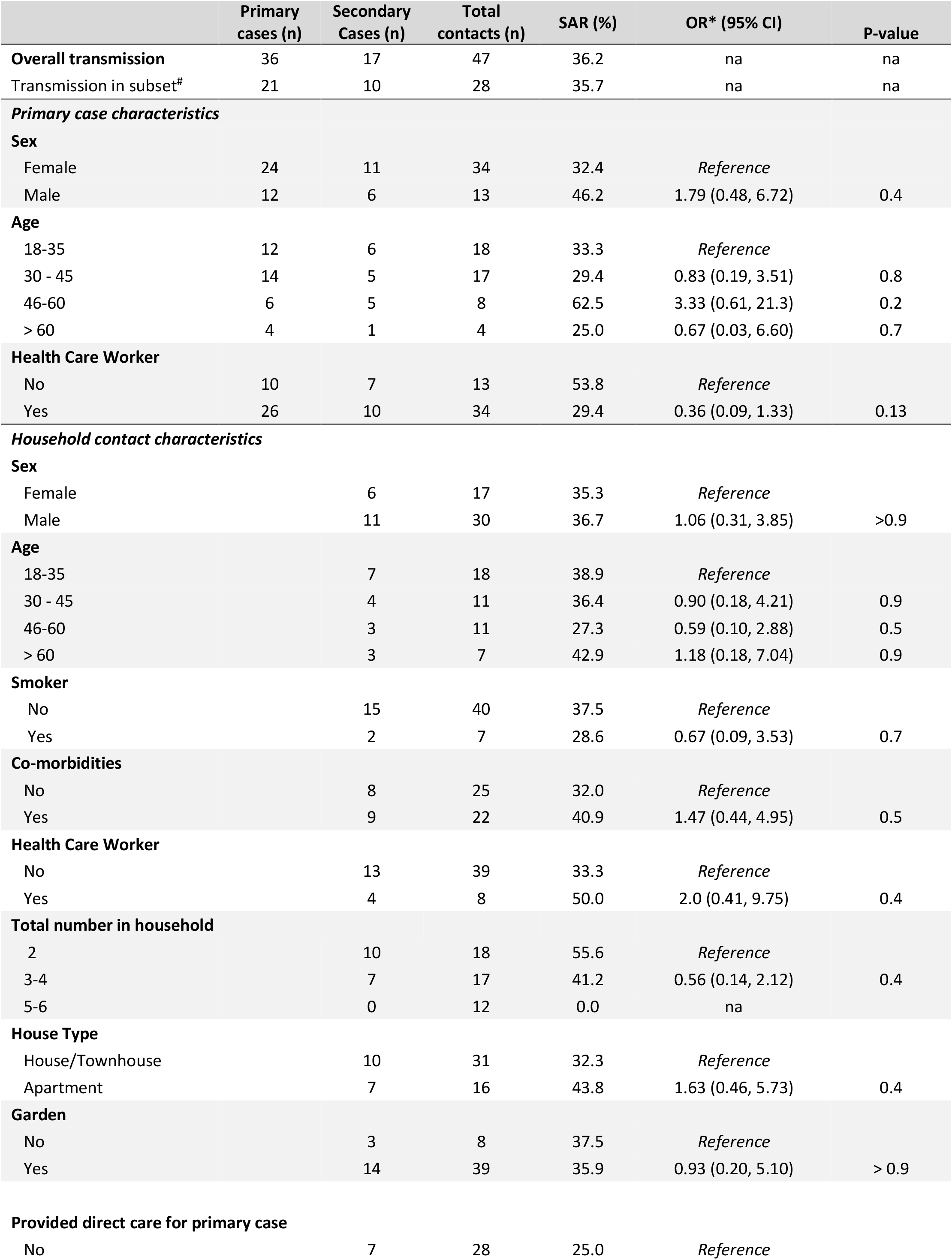
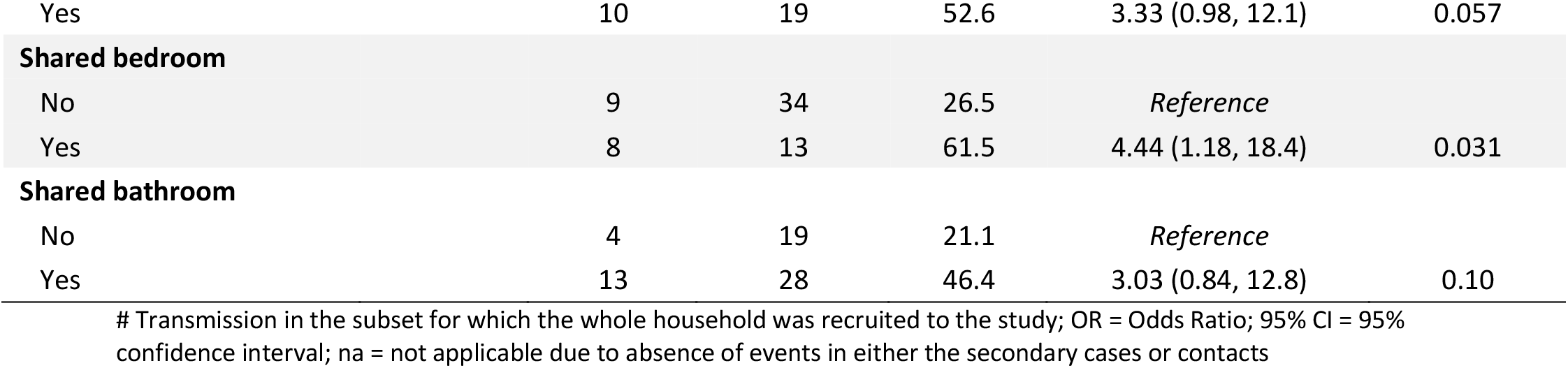
Secondary attack rates and odds ratio for transmission in household contacts.

### Population II) Health care workers in a hospital setting

A total of 1971 staff (~20% of all staff) participated in serosurvey 1, with 832 staff (~8% of all staff) participating across surveys 2 and 3. Staff sero-positivity rose from 0.05% (1/1971) in May 2020, to 3.8% (11/290) of those tested on Campus 2 in August and 5.5% (30/542) of those tested on Campus 1 in October (Table 2). All PCR tests in May were negative, except for one participant for which retrospective COVID-19 diagnosis was made by serology and whom had significant epidemiological risk factors for overseas acquisition in April. In August and October four new COVID-19 diagnoses were made by serology for staff at Campus 2 and one for staff at Campus 1, all had been working on COVID-19 wards. Four of the 5 described a symptomatic illness that they attributed to COVID-19, but did not experience respiratory symptoms. Symptoms included non-localising fever, migraine, diarrhoea, and fatigue. All had negative COVID-19 PCR swabs at the time of their symptomatic illness, as well as at other times in the preceding 6 months.

**Table 2:** COVID-19 Cross-sectional serosurvey results, RMH.

|  | Post Wave 1<br>(May 2020,<br>Campus 1) | Post Wave 2<br>(Aug 2020; Campus 2<br>Outbreak) | Post Wave 2<br>(Oct 2020; Campus 1,<br>HCA clusters) |
| --- | --- | --- | --- |
| Staff tested (n) | 1972 | 290 | 542 |
| PCR tests per<br>participant, n (IQR) | NA | 3 (2-5) | 2 (1-3) |
| Positive staff<br>confirmed by IgG (n) | 1 (0.05%) | 11 (3.8%) | 30 (5.5%) |
| New Dx by IgG | 1 | 4 (36.3%) | 1 (3.3%) |
| Sero-prevalence by<br>DHHS case count | 0.03% | 0.4% | 0.4% |
| PCR positive cases<br>with negative IgG | 0 | 2 | 0 |

OB = Outbreak; HCA = Health Care associated; Dx = diagnosis; DHHS = Department of Health and Human Services; IQR = Inter-quartile Range

Of the 832 participants who underwent serological testing, 701 had completed survey questions (44 COVID-19 positive staff, 657 COVID-19 negative staff) for which descriptive statistics are presented in Supplementary Table 3. No significant increase in sero-positivity was noted for those who described contact with a COVID-19 case unrelated to the hospital setting (7.6% versus 114%, p = 0.38)) or those exposed to an aerosol generating procedure (AGP) (40.3% versus 35.3%, p = 0.74). Of those variables which showed an increased rate of sero-positivity, multivariate analysis demonstrated that male HCW were less likely to be sero-positive (OR 0.19 (0.03-0.7), while nursing staff, staff who experienced anosmia and those caring for COVID-19 patients were more likely to be sero-positive (OR 8.1 (1.7-145.6) p 0.04; OR 22.7 (5.4-112.4) p < 0.001; OR 4.1 (1.8-10.5) p 0.01, respectively) (Table 3).

**Table 3:** HCW sero-survey results of univariate and multivariate analyses. Odds ratios and 95% confidence intervals are presented.

| Variable | Univariate |  | Multivariate |  |
| --- | --- | --- | --- | --- |
|  | OR (95% CI) | P-value | OR (95% CI) | P-value |
| **Sex** |  |  |  |  |
| Female | Reference |  | Reference |  |
| Male | 0.14 (0.02, 0.5) | 0.007 | 0.19 (0.03, 0.7) | 0.04 |
| **Role** |  |  |  |  |
| Allied health | Reference |  | Reference |  |
| Medical | 3.7 (0.5, 76.2) | 0.3 | 2.3 (0.2, 51.6) | 0.5 |
| Nursing | 13.1 (2.8, 234.7) | 0.01 | 8.1 (1.7, 145.6) | 0.04 |
| Other | 1.1 (0.1, 24.8) | 0.9 | 1.8 (0.1, 43.6) | 0.7 |
| **Direct patient care** |  |  |  |  |
| <10 hours | Reference |  | Reference |  |
| 10-20 hours | 2.6 (0.5, 10.9) | 0.2 | 0.9 (0.1, 5.5) | 0.9 |
| 20-50 hours | 3.8 (1.4, 12.2) | 0.01 | 1.3 (0.4, 5.2) | 0.7 |
| 50-80 hours | 5.5 (2.2, 16.8) | <0.001 | 1.5 (0.5, 5.8) | 0.5 |
| >80 hours | 1.7 (0.1, 10.9) | 0.6 | 1.4 (0.06, 11.2) | 0.8 |
| **Fever** |  |  |  |  |
| No | Reference |  | Reference |  |
| Yes | 4.3 (2, 8.4) | <0.001 | 1.8 (0.6, 4.8) | 0.3 |
| **Anosmia** |  |  |  |  |
| No | Reference |  | Reference |  |
| Yes | 21.2 (8, 56.7) | <0.001 | 22.7 (5.4, 112.4) | <0.001 |
| **Contact with COVID-19 patient** |  |  |  |  |
| No | Reference |  | Reference |  |
| Yes | 5.4 (2.7, 11.8) | <0.001 | 4.1 (1.8, 10.5) | 0.001 |

Table footnotes:
- Allied health includes: allied health assistant, physiotherapy, occupational therapy, podiatry, dietetics, social work, speech pathology, allied health students, phlebotomists, pharmacist - Medical includes: Consultants, Fellows, HMOs and medical students - Nursing includes: RN1, RN2, education nurse, psychiatry nurse, nursing student, other nurse - Other includes: Environmental services, food services, facilities, ward clerk, laboratory, admin

## Discussion

Rapid deployment of serological based testing has facilitated an improved understanding of COVID-19 transmission dynamics in this local setting. Serology demonstrated no additional benefit to the PCR testing of symptomatic household contacts for the detection of secondary cases of COVID-19 infection during mandatory home quarantine, however was helpful in finding additional COVID-19 cases in a health care setting with a high burden of infections. A rise in sero-prevalence for health care workers from 0.5% to ~5% was seen across this study period, making HCW more than 10 times more likely than the general population to become infected with COVID-19. In addition, the finding that caring for a COVID-19 patient was significantly associated with a greater likelihood of becoming sero-positive for COVID-19 (OR 4.1 (1.8-10.5), p 0.001) suggests that infection control measures were inadequate for staff during the second wave of COVID-19 infections in Victoria.

No asymptomatic infections were detected in this household cohort study. This is somewhat unexpected, as asymptomatic infections have been reported at rates of 17-40%, although the differentiation between very mild or pre-symptomatic cases is not always clear (14,15). There is the potential that some asymptomatic cases failed to seroconvert, and were thus missed given lower SARS-CoV-2 antibody seroconversion rates for asymptomatic (~60-80%), compared to mild (~80-95%) or severe infections (~98-100%)(16–19). Nevertheless, with the expected asymptomatic infection rates and seroconversion rates described here, some asymptomatic case detection would have been expected in this study group. This suggests for this cohort, with almost daily telephone contact with public health officers, the threshold for PCR testing was very low and detection rate high. With removal of barriers to access a test and any mild change from baseline health considered a symptom, all contacts were diagnosed by PCR. We do not have information on the number of PCR tests each contact underwent while in quarantine; if multiple tests were undertaken this would also reduce the likelihood of missing a COVID-19 diagnosis. A meta-analysis demonstrated that repeated PCR testing increased the detection of secondary cases (from 9.2% with one swab, to 21.2% with > 2 swabs) (20), and when multiple sample types are tested for SARS-CoV-2 the SAR for adults reached 51% in one study (21). In this cohort of relatively well household contacts, serological testing diagnosed fewer cases of COVID-19 infection than close monitoring of cases with symptom directed PCR testing (39/42 versus 42/42). Despite concerns regarding the transmission risk of asymptomatic infections, this study provides reassurance for local public health units that the approach at that time of only testing symptomatic contacts did not miss a significant number of transmission events.

The overall secondary attack rate (SAR) in this study, 36.2% (95% CI: 37.4-74.5%), was higher than both that of a retrospective NSW study including 229 households with 88 contacts and an Australian multi-state prospective study with 96 households and 286 contacts conducted at a similar period in the pandemic, at 22.5% and 15% respectively (22,23). It is also higher than the SAR of 17.1-18.9% found in two meta-analyses (84 and 2 studies, respectively)(20,24). As the primary aim of this study was to rapidly deploy serology to understand how many transmission events may be missed with PCR alone, rather than to determine SAR as a primary outcome, caution is advised in interpreting the SAR. Bias in recruiting only adults, as well as a higher proportion of health care workers, some of whom likely had occupational exposure to COVID-19 (while wearing PPE) prior to quarantine, means that despite using a more conservative definition for secondary cases (symptom onset at least 2 days apart) compared to other studies (symptom onset at least 1 day apart)(20,24), some secondary cases may have actually represented co-primary cases. In this study, an increased but not significant OR of 2.0 (0.41-9.75), for household contacts who were also HCW to acquire COVID-19, would be in keeping with this observation. Recall bias due to retrospective recollection of symptom onset may have led to inaccuracies in timing the onset of infection for some secondary cases, although most cases described onset on the same day as sample collection for PCR testing given these were the public health instructions at the time, the date of which was verified by investigators. Additional household and case/contact variables which were found on meta-analysis to be associated with a higher SAR and which may be contributory here include a high proportion of symptomatic cases (SAR 18.0% vs 0.7%), adult rather than child contacts (SAR 28% vs 16.8%), and smaller household numbers (1 contact versus 3 or more, SAR 41.5% vs 22.8%)(25). In this study many variables associated with an increased burden of contact between primary cases and household contacts (e.g. shared or smaller living spaces) demonstrated an elevated OR for COVID-19 transmission, although only sharing a bedroom was statistically significant. This study may have been underpowered to demonstrate the significance of these associations, as recruitment was limited by the number of cases of COVID-19 at this time of public lockdown measures.

Health care worker sero-prevalence studies have returned quite heterogenous results depending on the geographical area and study setting. Local factors such as community prevalence rates, local infection control practices (e.g. access to and utilisation of PPE for HCW, including N95 versus surgical respirator use), as well as the burden of COVID-19 admissions per unit of physical area (e.g. single rooms, negative pressure rooms versus cohorting) and ventilation and ward design of the health care setting are likely to influence COVID-19 transmission to HCW, although are not always well described in sero-survey reports. Three meta-analyses undertaken prior to vaccine availability report HCW sero-positivity rates of 7-9% (26–28), which are generally at least twice that of community prevalence (29,30). ‘High exposure’ to COVID-19 cases has been associated with increased sero-positivity rates (28), and in significant outbreak situations HCW sero-positivity has been reported in up to 30-40% (e.g. London(31,32), New York City(33,34)). In contrast, studies which have shown low levels of HCW sero-positivity are associated with low community prevalence and high access to PPE, including N95 respirators (35–37). Our finding of ~4-5% HCW sero-positivity is thus below the global average, but reflects an approximately 10 fold increase compared to the Australia community at the time(38), and twice that reported by another Victorian health service(39). The significantly increased rate of HCW infection at RMH compared to other local health services may relate to an increased burden of infection, as RMH was a hospital initially delegated to preferentially accept COVID-19 cases, and received a high number of residential aged care transfers, both of which were found to be risk factors for hospital acquired infections in a local Victorian study(4,40). Physical and engineering factors may also contribute to poor COVID-19 clearance and an increased risk of transmission, although higher rates were seen across both RMH campuses which encompassed different environments. Infection prevention measures were guided by public health recommendations, and thus were likely similar across both institutions prior to RMH moving to N95 respirators on 21 July, due to increased case numbers across the organisation.

Our findings of higher rates of infection in females, nursing staff and in those who experience anosmia are in keeping with published meta-analyses(26–28). Direct contact with COVID-19 has inconsistently been associated with higher sero-positivity rates, which likely reflects different transmission risks depending on the burden of cases, access to and type of PPE and PCR testing(28). The approximately 10 fold higher sero-positivity rate seen in our HCW cohort compared to the community, in addition to significantly increased OR for infection (4.1 (1.8-10.5), p 0.001) in those staff directly caring for COVID-19 patients, suggests that infection preventative actions were insufficient in this healthcare setting. Interestingly, AGP was not associated with a greater likelihood of COVID-19 infection in this study, suggesting that the enhanced PPE (N95 respirator use rather than surgical mask) and physical environment of the AGP (often single or negative pressure room) was successful in mitigating transmission risks. These findings are supportive of the early switch RMH made to utilising N95 respirator in all clinical settings managing COVID-19 infection, which was not stipulated by state public health guidelines at the time.

In this setting of frequent and rapid access to PCR testing, serological testing offered a small advantage in identifying additional staff infections, with 5 additional infections detected here (~12% of sero-positive cases) similar to other Australian studies (3/16 cases, ~ 18%)(39). A higher proportion of asymptomatic or false negative PCR results was seen in the context of a large outbreak (Campus 2, 4/11 (36%)), rather than in the environment of sustained smaller clusters (Campus 1, 1/30 (3.3%)) despite a higher sero-positivity rate at this site (3.8% vs 5.5%). Given that the 5 staff in this study reported an absence of respiratory symptoms, it is plausible that false negative results were related to low levels or absence of virus was present in the respiratory tract at time of testing rather than PCR analytical issues. In settings with lower access to high quality rapid PCR testing, serological testing provides a more significant advantage to case finding, with up to 40% additional cases detected(26,41–43).

This study is limited by being from a single site, with recruitment during a unique period of the pandemic in Victoria, Australia (May – October 2020). Staff testing numbers represent a small proportion of the available cohort (~8-20%), and there was likely some selection bias in a higher number of staff who knew they had been infected with COVID-19 presenting for testing (36 of 262 PCR reported staff infections, ~14%). A small number of other countries pursued a COVID-19 zero transmission for which these findings are generalisable, however as many countries moved towards COVID-19 endemicity some of the findings or public health approaches outlined here may be less relevant. However it is this unique environment of low community transmission but a high in-hospital event rate, that gives more significance to our HCW sero-survey results. Other strengths include robust serological and PCR testing and rapid mobilisation of testing to capture information about this window in the pandemic progression in Victoria.

## Conclusion

The rapid implementation of early serological assays added incrementally to that of regular PCR testing to provide real-time feedback on local infection control policies. Initiation of PCR testing for symptomatic household contacts was demonstrated to be a robust policy to detect COVID-19 transmission within a household, with the likelihood that few secondary cases are missed. Sero-surveys demonstrated increased sero-positivity in health care workers following a higher burden of care for COVID-19 patients, risk analysis suggested a failure of adequate infection prevention measures in some hospital COVID-19 wards at that time.

## Data Availability

All data produced in the present study are available upon reasonable request to the authors

## Acknowledgments

We thank the scientific staff at the Pathology Department; the staff of the Infection Prevention and Surveillance Service and the staff of RMH@Home at the Royal Melbourne Hospital; as well as the staff the Serology Department of the Victorian Infection Diseases Reference Laboratory. We thank Dr. Martin Dutch for his support with REDCap and Prof. Jo Douglass for support from the RMH Office for Research. Finally we would like to thank all RMH staff who participated in these sero-surverys and the COVID-19 patients and their households who participated in the household sero-surveys.

## References

1. Australian Health Protection Principal Committee (AHPPC) statement on strategic direction _ Australian Government Department of Health and Aged Care.pdf [Internet]. Australian Government Department of Health and Aged Care; Available from: https://www.health.gov.au/news/australian-health-protection-principal-committee-ahppc-statement-on-strategic-direction

2. Victorian Department of Health COVID-19 writing group, group. Population-based analysis of the epidemiological features of COVID-19 epidemics in Victoria, Australia, January 2020 - March 2021, and their suppression through comprehensive control strategies. The Lancet Regional Health - Western Pacific. 2021;17.

3. Lane CR, Sherry NL, Porter AF, Duchene S, Horan K, Andersson P, et al. Genomics-informed responses in the elimination of COVID-19 in Victoria, Australia: an observational, genomic epidemiological study. The Lancet Public Health. 2021 Aug;6(8):e547–56.

4. Buising KL, Williamson D, Cowie BC, MacLachlan J, Orr E, MacIsaac C, et al. A hospital-wide response to multiple outbreaks of COVID-19 in health care workers: lessons learned from the field. Medical Journal of Australia. 2021 Feb;214(3):101.

5. World Health Organization (Global). Clinical management of COVID-19 [Internet]. 2020. Available from: https://www.who.int/publications/i/item/clinical-management-of-covid-19

6. Corman VM, Landt O, Kaiser M, Molenkamp R, Meijer A, Chu DK, et al. Detection of 2019 novel coronavirus (2019-nCoV) by real-time RT-PCR. Eurosurveillance [Internet]. 2020 Jan 23 [cited 2020 Mar 25];25(3). Available from: https://www.eurosurveillance.org/content/10.2807/1560-7917.ES.2020.25.3.2000045

7. Williams E, Bond K, Chong B, Giltrap D, Eaton M, Kyriakou P, et al. Implementation and evaluation of a novel real-time multiplex assay for SARS-CoV-2: in-field learnings from a clinical microbiology laboratory. Pathology. 2020 Dec;52(7):754–9.

8. Loeffelholz MJ, Alland D, Butler-Wu SM, Pandey U, Perno CF, Nava A, et al. Multicenter Evaluation of the Cepheid Xpert Xpress SARS-CoV-2 Test. McAdam AJ, editor. J Clin Microbiol. 2020 May 4;58(8):e00926-20, /jcm/58/8/JCM.00926-20.atom.

9. Abbott. Alinity m SARS-CoV-2 assay - instructions for use [Internet]. [cited 2021 Dec 20]. Available from: https://www.fda.gov/media/137979/download

10. DiaSorin Liaison SARS-CoV-2 S1/S2 IgG, Ref 311450, EN – 200/007-797, 07-20202-07.

11. Bond KA, Williams E, Nicholson S, Lim S, Johnson D, Cox B, et al. Longitudinal evaluation of laboratory-based serological assays for SARS-CoV-2 antibody detection. Pathology. 2021 Oct;53(6):773–9.

12. GenScript SARS-CoV-2 Surrogate Virus Neutralization Test Kit instructions for use L00847-5, Version 3.1 update 04/29/2020.

13. Harris PA, Taylor R, Thielke R, Payne J, Gonzalez N, Conde JG. Research electronic data capture (REDCap)—A metadata-driven methodology and workflow process for providing translational research informatics support. Journal of Biomedical Informatics. 2009 Apr;42(2):377–81.

14. Byambasuren O, Cardona M, Bell K, Clark J, McLaws ML, Glasziou P. Estimating the extent of asymptomatic COVID-19 and its potential for community transmission: Systematic review and meta-analysis. Official Journal of the Association of Medical Microbiology and Infectious Disease Canada. 2020 Dec;5(4):223–34.

15. Ma Q, Liu J, Liu Q, Kang L, Liu R, Jing W, et al. Global Percentage of Asymptomatic SARS-CoV-2 Infections Among the Tested Population and Individuals With Confirmed COVID-19 Diagnosis: A Systematic Review and Meta-analysis. JAMA Netw Open. 2021 Dec 14;4(12):e2137257.

16. Rijkers G, Murk JL, Wintermans B, Van Looy B, Van Den Berge M, Veenemans J, et al. Differences in Antibody Kinetics and Functionality Between Severe and Mild Severe Acute Respiratory Syndrome Coronavirus 2 Infections. The Journal of Infectious Diseases. 2020 Sep 14;222(8):1265–9.

17. Ko JH, Joo EJ, Park SJ, Baek JY, Kim WD, Jee J, et al. Neutralizing Antibody Production in Asymptomatic and Mild COVID-19 Patients, in Comparison with Pneumonic COVID-19 Patients. JCM. 2020 Jul 17;9(7):2268.

18. Lei Q, Li Y, Hou H, Wang F, Ouyang Z, Zhang Y, et al. Antibody dynamics to SARS-CoV-2 in asymptomatic COVID-19 infections. Allergy. 2021 Feb;76(2):551–61.

19. Milani GP, Dioni L, Favero C, Cantone L, Macchi C, Delbue S, et al. Serological follow-up of SARS-CoV-2 asymptomatic subjects. Sci Rep. 2020 Nov 18;10(1):20048.

20. Fung HF, Martinez L, Alarid-Escudero F, Salomon J, Studdert D, Andrews J, et al. The household secondary attack rate of SARS-CoV-2: A rapid review. 2020; Available from: https://semanticscholar.org/paper/664dd153156e924bc49bdd08a39a34ccec738005

21. Reukers DFM, Van Boven M, Meijer A, Rots N, Reusken C, Roof I, et al. High Infection Secondary Attack Rates of Severe Acute Respiratory Syndrome Coronavirus 2 in Dutch Households Revealed by Dense Sampling. Clinical Infectious Diseases. 2022 Jan 7;74(1):52–8.

22. Sordo AA, Dunn A, Gardiner ER, Reinten TA, Tsang TS, Deng L, et al. Household transmission of COVID-19 in 2020 in New South Wales, Australia. Commun Dis Intell (2018) [Internet]. 2022 Apr 26 [cited 2024 Jun 23];46. Available from:https://www1.health.gov.au/internet/main/publishing.nsf/Content/2A15CD097063EF40CA2587CE008354F1/$File/household_transmission_of_covid_19_in_2020_in_new_south_wales_australia.pdf

23. Marcato AJ, Black AJ, Walker CR, Morris D, Meagher N, Price DJ, et al. Learnings from the Australian first few X household transmission project for COVID-19. The Lancet Regional Health - Western Pacific. 2022 Nov;28:100573.

24. Madewell ZJ, Yang Y, Longini IM, Halloran ME, Dean NE. Factors Associated With Household Transmission of SARS-CoV-2: An Updated Systematic Review and Meta-analysis. JAMA Netw Open. 2021 Aug 27;4(8):e2122240.

25. Madewell ZJ, Yang Y, Longini IM, Halloran ME, Dean NE. Household Transmission of SARS-CoV-2: A Systematic Review and Meta-analysis. JAMA Netw Open. 2020 Dec 14;3(12):e2031756.

26. Gómez-Ochoa SA, Franco OH, Rojas LZ, Raguindin PF, Roa-Díaz ZM, Wyssmann BM, et al. COVID-19 in Health-Care Workers: A Living Systematic Review and Meta-Analysis of Prevalence, Risk Factors, Clinical Characteristics, and Outcomes. American Journal of Epidemiology. 2021 Jan 4;190(1):161–75.

27. Galanis P, Vraka I, Fragkou D, Bilali A, Kaitelidou D. Seroprevalence of SARS-CoV-2 antibodies and associated factors in healthcare workers: a systematic review and meta-analysis. Journal of Hospital Infection. 2021 Feb;108:120–34.

28. Kayı i, Madran B, Keske Ş, Karanfil Ö, Arribas JR, Pshenichnaya N, et al. The seroprevalence of SARS-CoV-2 antibodies among health care workers before the era of vaccination: a systematic review and meta-analysis. Clinical Microbiology and Infection. 2021 Sep;27(9):1242–9.

29. Rostami A, Sepidarkish M, Leeflang MMG, Riahi SM, Nourollahpour Shiadeh M, Esfandyari S, et al. SARS-CoV-2 seroprevalence worldwide: a systematic review and meta-analysis. Clinical Microbiology and Infection. 2021 Mar;27(3):331–40.

30. Ebinger JE, Botwin GJ, Albert CM, Alotaibi M, Arditi M, Berg AH, et al. Seroprevalence of antibodies to SARS-CoV-2 in healthcare workers: a cross-sectional study. BMJ Open. 2021 Feb;11(2):e043584.

31. Grant JJ, Wilmore SMS, McCann NS, Donnelly O, Lai RWL, Kinsella MJ, et al. Seroprevalence of SARS-CoV-2 antibodies in healthcare workers at a London NHS Trust. Infect Control Hosp Epidemiol. 2020 Aug 4;1–3.

32. Houlihan CF, Vora N, Byrne T, Lewer D, Kelly G, Heaney J, et al. Pandemic peak SARS-CoV-2 infection and seroconversion rates in London frontline health-care workers. The Lancet. 2020 Jul;396(10246):e6–7.

33. Mansour M, Leven E, Muellers K, Stone K, Mendu DR, Wajnberg A. Prevalence of SARS-CoV-2 Antibodies Among Healthcare Workers at a Tertiary Academic Hospital in New York City. J GEN INTERN MED. 2020 Aug;35(8):2485–6.

34. Venugopal U, Jilani N, Rabah S, Shariff MA, Jawed M, Mendez Batres A, et al. SARS-CoV-2 seroprevalence among health care workers in a New York City hospital: A cross-sectional analysis during the COVID-19 pandemic. International Journal of Infectious Diseases. 2021 Jan;102:63–9.

35. Ko JH, Lee JY, Kim HA, Kang SJ, Baek JY, Park SJ, et al. Serologic Evaluation of Healthcare Workers Caring for COVID-19 Patients in the Republic of Korea. Front Microbiol. 2020 Nov 20;11:587613.

36. Woon YL, Lee YL, Chong YM, Ayub NA, Krishnabahawan SL, Lau JFW, et al. Serology surveillance of SARS-CoV-2 antibodies among healthcare workers in COVID-19 designated facilities in Malaysia. The Lancet Regional Health - Western Pacific. 2021 Apr;9:100123.

37. Korth J, Wilde B, Dolff S, Anastasiou OE, Krawczyk A, Jahn M, et al. SARS-CoV-2-specific antibody detection in healthcare workers in Germany with direct contact to COVID-19 patients. Journal of Clinical Virology. 2020 Jul;128:104437.

38. Vette KM, Machalek DA, Gidding HF, Nicholson S, O’Sullivan MVN, Carlin JB, et al. Seroprevalence of Severe Acute Respiratory Syndrome Coronavirus 2-Specific Antibodies in Australia After the First Epidemic Wave in 2020: A National Survey. Open Forum Infectious Diseases. 2022 Mar 1;9(3):ofac002.

39. Lau JS, Buntine P, Price M, Darzins P, Newnham E, Connell A, et al. SARS-CoV-2 seroprevalence in healthcare workers in a tertiary healthcare network in Victoria, Australia. Infection, Disease & Health. 2021 Aug;26(3):208–13.

40. Victorian Government. COVID-19 Hospital-acquired infections among patients in Victorian Health Services (25 January 2020 - 15 November 2020) [Internet]. 2021. Available from: https://nla.gov.au/nla.obj-3058817726/view

41. Nakagama Y, Komase Y, Candray K, Nakagama S, Sano F, Tsuchida T, et al. Serological Testing Reveals the Hidden COVID-19 Burden among Health Care Workers Experiencing a SARS-CoV-2 Nosocomial Outbreak. Long SW, editor. Microbiol Spectr. 2021 Oct 31;9(2):e01082–21.

42. Ludewick H, Hahn R, Italiano C, Pereira L, Fatovich D, Saxton J, et al. COVID-19 Serosurvey of Frontline Healthcare Workers in Western Australia. J Epidemiol Glob Health. 2022 Sep 21;12(4):472–7.

43. Garcia-Basteiro AL, Moncunill G, Tortajada M, Vidal M, Guinovart C, Jiménez A, et al. Seroprevalence of antibodies against SARS-CoV-2 among health care workers in a large Spanish reference hospital. Nat Commun. 2020 Jul 8;11(1):3500.

