## Supplementary data for "Applying SARS-CoV-2 serological testing to understand the effectiveness of local infection control measures for COVID-19 infection in Victoria, Australia 2020"

**Supplementary Figure 1**

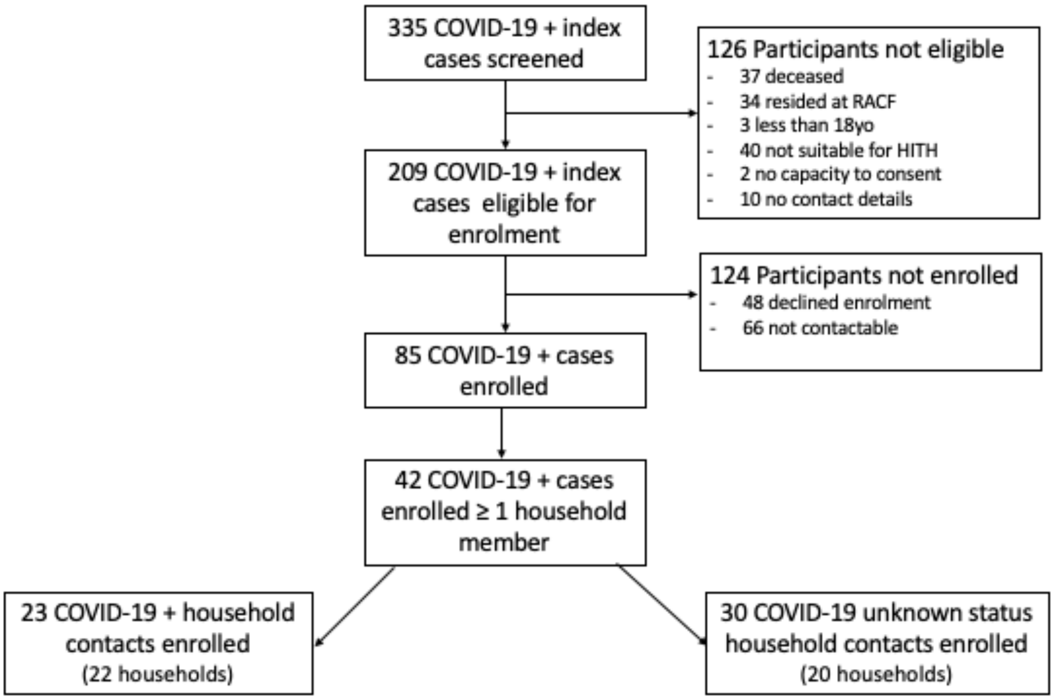

### Supplementary Table 1: Baseline participant characteristics

| | All index participants in primary study [*n* = 81] (%) | Index participants included in household analysis [*n* = 42] (%) | Household contacts with confirmed COVID-19 infection [*n* = 23] (%) | Household contacts with unknown COVID-19 status [*n* = 30] (%) | p value* |
|---|---|---|---|---|---|
| **Female** | 55 (67.9) | 26 (61.9) | 9 (39.1) | 11 (36.7) | 0.64 |
| **Age, median, IQR, (range)** | 34, 27-52 (22, 93) | 34, 27-49 (22, 91) | 37, 26-44 (21, 82) | 34, 28-56 (19, 77) | 0.7 |
| **Born in Australia** | 50 (61.7) | 27 (64.3) | 10 (43.5) | 19 (63.3) | 0.2 |
| **Primary language English** | 72 (88.9) | 39 (92.9) | 20 (87.0) | 25 (83.3) | 0.4 |
| **Employment Status** | | | | | 0.075 |
| *Full Time* | 36 (44.4) | 17 (40.5) | 15 (65.2) | 18 (60.0) | |
| *Part Time* | 37 (45.7) | 19 (45.2) | 3 (13.0) | 7 (23.3) | |
| *Other* | 9 (11.1) | 6 (14.3) | 5 (21.7) | 5 (16.7) | |
| **Health Care Worker** | 66 (81.5) | 31 (73.8) | 6 (26.1) | 4 (13.3) | < 0.001 |
| **Current smoker** | 3 (3.7) | 1 (2.4) | 2 (8.7) | 5 (16.7) | 0.1 |
| **Medical Co-morbidities** | 34 (42.0) | 17(40.5) | 9 (39.1) | 13 (40) | > 0.9 |
| **Experienced symptoms** | 78 (96.3) | 34 (95.2) | 22 (95.7) | 1 (3.3) | < 0.001 |
| **Severity of illness** | | | | n/a | < 0.001 |
| *Mild* | 74 (91.4) | 37 (88.1) | 23 (100) | | |
| *Moderate* | 5 (6.2) | 4 (9.5) | 0 (0) | | |
| *Severe* | 2 (2.5) | 1 (2.4) | 0 (0) | | |
| **Admitted to Hospital** | 17 () | 5 (11.9) | 1 (4.3) | n/a | < 0.001 |
| **No. of household members recruited** | na | | | | 0.004 |
| *2* | | 36 (85.7) | 20 (87) | 15 (50) | |
| *3-4* | | 5 (11.9) | 3 (13) | 10 (33.3) | |
| *5-6* | | 1 (2.4) | 0 (0) | 2 (16.7) | |
| **Total No. of Household** | na | | | | 0.011 |
| *2* | | 22 (52.4) | 14 (60.9) | 8 (26.7) | |
| *2-4* | | 14 (33.3) | 8 (34.8) | 10 (33.3) | |
| *5-6* | | 6 (14.3) | 1 (4.3) | 12 (40) | |
| **No. of Households with all members recruited** | na | 25 (59.5) | 14 (60.9) | 18 (60) | > 0.9 |
| **Relationship between household members** | na | | | | 0.2 |
| *Partners* | | 27 (64.3) | 15 (65.2) | 12 (40) | |
| *Family* | | 9 (21.4) | 5 (21.7) | 13 (43.3) | |
| *Not Related* | | 6 (14.3) | 3 (13) | 5 (16.7) | |
| **Household Age Group** | na | | | | 0.8 |
| *Adults* | | 28 (66.7) | 18 (78.3) | 19 (63.3) | |
| *Adults & Children* | | 7 (16.7) | 2 (8.7) | 6 (20) | |
| *Adults & Older Adults#* | | 7 (16.7) | 3 (13) | 5 (16.7) | |

*For recruited participants, i.e. Not including All index participants; # Older adults = > 60yrs

**Supplementary Table 2: Household transmission results of univariate and multivariate analysis**

| Variable | Univariate | | Multivariate | |
|---|---|---|---|---|
| | OR (95% CI) | P-value | OR (95% CI) | P-value |
| **Primary case** | | | | |
| *Health care worker* | | | | |
| No | *Reference* | | *Reference* | |
| Yes | 0.36 (0.09, 1.33) | 0.13 | 0.23 (0.04, 1.21) | 0.09 |
| **Secondary case** | | | | |
| *Provided direct care for PC* | | | | |
| No | *Reference* | | *Reference* | |
| Yes | 3.33 (0.98, 12.1) | 0.057 | 2.76 (0.65, 13.1) | 0.2 |
| *Shared a bedroom* | | | | |
| No | *Reference* | | *Reference* | |
| Yes | 4.44 (1.18, 18.4) | 0.031 | 7.46 (1.34, 53.3) | 0.029 |
| *Shared a bathroom* | | | | |
| No | *Reference* | | *Reference* | |
| Yes | 3.03 (0.84, 12.8) | 0.10 | 1.72 (0.35, 9.16) | 0.5 |

OR = Odds Ratio; 95% CI = 95% confidence interval; HCW – Health Care Worker; PC = Primary case

**Supplementary Table 3. Health Care Worker participant characteristics and exposures**

| | Negative n=657 | Positive n=44 | Total n=701 | P-value |
|---|---|---|---|---|
| **DEMOGRAPHICS** | | | | |
| *Gender* | | | | |
| Female | 492 (74.9%) | 42 (95.5%) | 534 (76%) | 0.002 |
| Male | 162 (24.7%) | 2 (4.5%) | 164 (23%) | |
| *Age group* | | | | |
| 18-24 years | 45 (6.8%) | 4 (9.1%) | 49 (7%) | 0.56 |
| 25-44 years | 339 (51.6%) | 27 (61.4%) | 366 (52%) | |
| 45-64 years | 239 (36.4%) | 13 (29.5%) | 252 (36%) | |
| 65+ years | 19 (2.9%) | - | 19 (3%) | |
| **EMPLOYMENT INFORMATION** | | | | |
| *Role* | | | | |
| Allied health | 102 (14.6%) | 1 (0.1%) | 103 (14.7%) | <0.001 |
| Medical | 82 (11.7%) | 3 (0.4%) | 85 (12.1%) | |
| Nursing | 295 (42.1%) | 38 (5.4%) | 333 (47.5%) | |
| Other | 178 (25.4) | 2 (0.3%) | 179 (25.5%) | |
| *Employment* | | | | |
| Full time | 258 (39.3%) | 10 (22.7%) | 268 (38%) | 0.05 |
| Part time | 365 (55.6%) | 29 (65.9%) | 394 (56%) | |
| Casual | 30 (4.6%) | 5 (11.4%) | 35 (5%) | |
| *Direct patient care* | | | | |
| <10 hours | 234 (35.6%) | 5 (11.4%) | 239 (34%) | 0.003 |
| 10-20 hours | 54 (8.2%) | 3 (6.8%) | 57 (8%) | |
| 20-50 hours | 147 (22.4%) | 12 (27.3%) | 159 (23%) | |
| 50-80 hours | 194 (29.5%) | 23 (52.3%) | 217 (31%) | |
| >80 hours | 28 (4.3%) | 1 (2.3%) | 29 (4%) | |
| Cared for COVID or SCOVID patients | 280 (42.6%) | 33 (75%) | 313 (45%) | <0.001 |
| Worked at another hospital | 95 (14.5%) | 5 (11.4%) | 100 (14%) | 0.82 |
| Worked at aged care facility | 13 (2%) | 2 (4.5%) | 15 (2%) | 0.24 |
| **COVID TESTING** | | | | |
| *Number of swabs for COVID-19* | | | | |
| None | 55 (8.4%) | 2 (4.5%) | 57 (8%) | 0.02 |
| 1-5 | 433 (65.9%) | 39 (88.6%) | 472 (67%) | |
| 6-9 | 111 (16.9%) | 2 (4.5%) | 113 (16%) | |
| >10 | 58 (8.8%) | 1 (2.3%) | 59 (8%) | |
| Had at least one illness compatible with COVID-19 | 192 (29.2%) | 19 (43.2%) | 211 (30%) | 0.06 |
| Fever | 59 (9%) | 13 (29.5%) | 72 (10%) | <0.001 |
| Anosmia | 9 (1.4%) | 10 (22.7%) | 19 (3%) | <0.001 |
| Contact with COVID-19 non-patient | 50 (7.6%) | 5 (11.4%) | 55 (8%) | 0.38 |
| **RISK FACTORS FOR COVID ACQUISITION** | | | | |
| Contact with COVID-19 patient | 253 (38.5%) | 34 (77.3%) | 287 (41%) | <0.001 |
| | n=253 | n=34 | n=287 | |
| Exposure to AGP* | 102 (40.3%) | 12 (35.3%) | 114 (40%) | 0.74 |
| Proximity to COVID patient during patient care* | | | | |
| Within 1.5 meters | 230 (90.9%) | 29 (85.3%) | 259 (90%) | 0.35 |
| Not within 1.5 meters | 20 (7.9%) | 4 (11.8%) | 24 (8%) | |
| COVID positive patient who was shouting, singing, coughing, or vomiting* | 191 (75.5%) | 30 (88.2%) | 221 (77%) | 0.13 |
| COVID positive patient who was confused, agitated, or wandering* | 119 (47%) | 26 (76.5%) | 145 (51%) | <0.001 |

*Variable only applies to those who reported contact with a COVID-19 patient. There were 27 missing responses across all datapoints, 26 from COVID-19 negative and 1 from a COVID-19 positive staff members. SCOVID = Suspected COVID-19 infection.
